# Seroconversion of a city: Longitudinal monitoring of SARS-CoV-2 seroprevalence in New York City

**DOI:** 10.1101/2020.06.28.20142190

**Authors:** Daniel Stadlbauer, Jessica Tan, Kaijun Jiang, Matthew M. Hernandez, Shelcie Fabre, Fatima Amanat, Catherine Teo, Guha Asthagiri Arunkumar, Meagan McMahon, Jeffrey Jhang, Michael D. Nowak, Viviana Simon, Emilia Mia Sordillo, Harm van Bakel, Florian Krammer

## Abstract

By conducting a retrospective, cross-sectional analysis of SARS-CoV-2 seroprevalence in a ‘sentinel group’ (enriched for SARS-CoV-2 infections) and a ‘screening group’ (representative of the general population) using >5,000 plasma samples from patients at Mount Sinai Hospital in New York City (NYC), we identified seropositive samples as early as in the week ending February 23, 2020. A stark increase in seropositivity in the sentinel group started the week ending March 22 and in the screening group in the week ending March 29. By the week ending April 19, the seroprevalence in the screening group reached 19.3%, which is well below the estimated 67% needed to achieve community immunity to SARS-CoV-2. These data potentially suggest an earlier than previously documented introduction of SARS-CoV-2 into the NYC metropolitan area.

**One Sentence Summary:** Seroprevalence of SARS-CoV-2 in cross-sectional samples from New York City rose from 0% to 19.3% from early February to mid-April.

## Main Text

The first case of Coronavirus Disease 2019 (COVID-19) was identified in NYC at Mount Sinai Hospital on February 29, 2020 (*1*). A sharp rise in severe acute respiratory syndrome coronavirus 2 (SARS-CoV-2) infections started to occur shortly afterwards during the week ending on March 8, followed by a significant increase of COVID-19 deaths during the week ending on March 15 (**Figure 1**). New York State implemented a stay-at-home order called the ‘New York on Pause Program’ effective at 8pm on March 22, 2020, and, as a consequence, daily case numbers in both New York State and NYC started to plateau and then decreased in April 2020. Although nucleic acid amplification testing (NAAT) is now widely available in New York State, there was little testing capacity at the beginning of the local epidemic in early March, and many mild to moderate cases likely went undetected. In addition, asymptomatic cases might have been missed since, in the absence of symptoms, NAAT would not have been recommended.

**Fig. 1.**
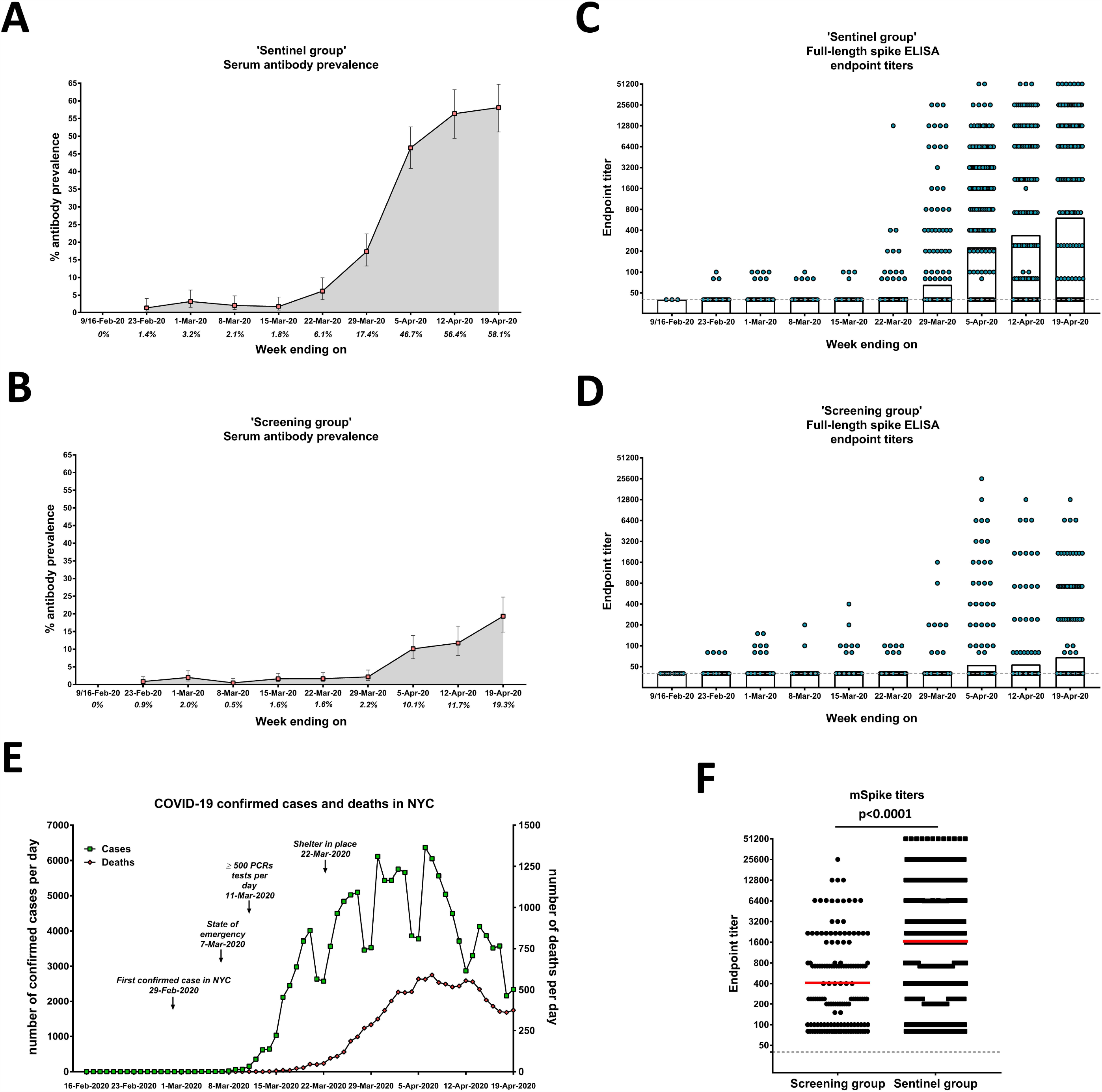
Seroprevalence, full-length spike antibody titers and confirmed cases in NYC. **(A)** Serum antibody prevalence in the sentinel group between the weeks ending February 9 (first two weeks combined) to April 19, 2020. **(B)** Serum antibody prevalence during that same timeframe in the screening group. **(C)** Full-length spike antibody titers in the sentinel group in the sampled time period. **(D)** Spike endpoint titers in the screening group. **(E)** Confirmed cases and deaths in NYC in the early weeks of the SARS-CoV-2 epidemic. **(F)** Comparison of endpoint titers in the screening and sentinel groups.

Although it is currently unknown if previous infection with SARS-CoV-2 can protect from reinfection, there are data from SARS-CoV-2 infection of non-human primates as well as from studies with other human coronaviruses suggesting that infection may confer immunity (*2, 3*). It is therefore important to determine the true infection rates in a population in order to assess how close this population is to potential ‘community immunity’ (*4*). Knowing the true infection rate also allows calculation of the infection fatality rate (IFR), which is very likely much lower than the case fatality rate (CFR). To estimate true infection rates, serosurveys can be used that measure the presence of antibodies that have been mounted to past virus infections, rather than the presence of virus. Several serological assays for measuring antibodies to SARS-CoV-2 have been developed (*5*). Many focus either on the virus nucleoprotein, the spike protein on the virus surface, or the receptor binding domain (RBD), which is an important part of the spike protein that interacts with angiotensin-converting enzyme 2 (ACE2), the cellular receptor for SARS-CoV-2 (*6*). We have recently established a two-step enzyme-linked immunosorbent assay (ELISA) in which serum/plasma samples are prescreened at a set dilution for reactivity to RBD. Positives in this first step are confirmed and the antibody titer assessed in a second step against a stabilized version of the full-length spike protein (*7, 8*). The use of two sequential assays reduces the false positive rate and favors high specificity. The assay used here has a workflow that closely resembles an assay established in the Mount Sinai Health System (MSHS) CLIA-certified Clinical Pathology Laboratory, which received an FDA emergency use authorization (EUA) for this ELISA in April 2020. However, the assay used in this study was performed in a research laboratory setting. An initial test of the assay performance with a panel of negative and positive samples suggested that the research-grade assay has a sensitivity of 95% and a specificity of 100% (**S. Table 1**). This results in a positive predictive value of 1 (PPV, 95% confidence interval (CI): 0.908-1.000) and a negative predictive value of 0.97 (NPV, 95% CI: 0.909-0.995). In the week of February 9, 2020, we started to collect random, de-identified, cross-sectional plasma samples and stored for standard of care medical purposes by the MSHS Clinical Pathology laboratories. These samples were divided into two distinct patient groups. The first group included samples from patients seen in Mount Sinai’s emergency department (ED) and from patients that were admitted to the hospital from the ED during the period beginning with the week ending on February 9 till the week ending on April 19. This group, termed the ‘sentinel group’, served as an indicator for SARS-CoV-2 infection since we assumed that individuals with moderate to severe COVID-19 would come to the ED and would be admitted to the hospital at increasing rates as the epidemic progressed. The second group of samples, termed the ‘screening group’, were obtained from patients at OB/GYN visits and deliveries, oncology-related visits, as well as hospitalizations due to elective or planned surgeries, transplant surgeries, pre-operative medical assessments and related outpatient visits, cardiology office visits, and other regular office/treatment visits. We reasoned that these samples might resemble more closely the general population because the purposes for these scheduled visits were unrelated to COVID-19. The sentinel group comprised 43.6% females while the screening group included 65.8% females (**Table 1**). The majority of individuals in the sentinel group were 61 years of age or older while the screening group had a more balanced age distribution resembling the general population (**Table 1**). Except for the weeks ending February 9 and February 16, for which only 16 samples were obtained across the two weeks (3 in the sentinel group, 13 in the screening group), the sentinel group size ranged between 195 and 274 samples per week and the screening group included 230-493 samples per week (**S. Table 2**). A total of 5,485 samples obtained from patients between the weeks ending February 9 and April 19 were tested: 3,412 samples in the screening group and 2,073 in the sentinel group. In the sentinel group, no positives were detected in the week ending February 9 and 16 and low seroprevalence was found between the weeks ending February 23 to March 15 (ranges between 1.4 and 3.2%, **Figure 1A**). While we believe these positives are true positives, the prevalence is low and within the confidence intervals of the PPV. A sharp increase to 6.1% was detected in the week ending March 22. This increase continued in the weeks ending March 29 (17.4%), April 5 (46.7%) and April 12 (56.4%); however, seropositivity in the sentinel group seemingly plateaued in the week ending April 19 at 58.1%. In the context of COVID-19 case and death rates reported for NYC, the seroprevalence values we report reflect hospital admissions due to COVID-19, although the uptick in positive serology results lagged approximately one to two weeks behind the increased molecular detection of SARS-CoV-2 infections. This is expected since there is usually a delay between infection and seroconversion. In summary, the numbers in the sentinel group are a reflection of hospital admissions due to COVID-19.

**Table 1:** Demographic characteristics, seroprevalence and COVID-19 diagnosis in different study populations.

| Population | n | Female | Age group |  |  |  | Antibody positive | COVID-19 diagnosis | Description |
| --- | --- | --- | --- | --- | --- | --- | --- | --- | --- |
|  |  |  | 0-20 | 21-40 | 41-60 | ≥61 |  |  |  |
| ‘Screening group’ |  |  |  |  |  |  |  |  |  |
| OB/GYN | 1366 | 1275<br>(93.3%) | 202<br>(14.8%) | 1054<br>(77.2%) | 96<br>(7.0%) | 14<br>(1.0%) | 81 (5.9%) | 49 (3.8%) | OB/GYN visits and deliveries |
| Oncology | 1319 | 614<br>(46.6%) | 46<br>(3.5%) | 212<br>(16.1%) | 342<br>(25.9%) | 719<br>(54.5%) | 42<br>(3.2%) | 35 (2.7%) | Oncology visits and treatment hospitalizations |
| Surgery | 544 | 263<br>(48.4%) | 13<br>(2.4%) | 66<br>(12.1%) | 190<br>(34.9%) | 277<br>(50.9%) | 17<br>(3.1%) | 16<br>(2.9%) | Surgeries, transplant surgeries, pre-op medical assessments and related visits |
| Cardiology and other office visits | 183 | 84<br>(45.9%) | 4 (2.2%) | 48<br>(26.2%) | 48<br>(26.2%) | 83<br>(45.4%) | 5<br>(2.7%) | 4<br>(2.2%) | Cardiology office visits and other regular office/treatment visits |
| Subtotal | 3412 | 2246<br>(65.8%) | 265<br>(7.8%) | 1380<br>(40.4%) | 674<br>(19.8%) | 1093<br>(32.0%) | 145<br>(4.3%) | 104<br>(3.1%) |  |
| ‘Sentinel group’ |  |  |  |  |  |  |  |  |  |
| Inpatient | 2073 | 903<br>(43.6%) | 57<br>(2.7%) | 273<br>(13.2%) | 608<br>(29.3%) | 1132<br>(54.6%) | 436<br>(21.0%) | 541<br>(26.1%) | Emergency department and inpatient admissions |

Similar to the sentinel group, the seroprevalence found in the screening group was very low during the weeks ending February 9 through March 29 (0% to 2%, **Figure 1B**). Of note, some samples during that time had moderately high reactivity (endpoint titers of 1:150-1:400) (**Figure 1D**). An increase in seroprevalence from 1.6% to 2.2% was detected in the week ending March 29, followed by increases to 10.1% and 11.7% in the following weeks, up to 19.3% seroprevalence in the week ending April 19. These numbers are significantly lower than the percentages calculated in the same weeks for the sentinel group, which makes sense since the visits from which the screening samples are derived are not related to COVID-19 infections and therefore are not enriched in SARS-CoV-2 infected patients. In addition, the delay between the sharp increase in SARS-CoV-2 detection by NAAT in NYC and the increase in seroprevalence in our screening group is longer than the delay between the increase in confirmed cases and the increase in seroprevalence in the sentinel group. This can be attributed to different antibody kinetics in mild cases, which likely constitute the majority of infections in the screening group. We have recently shown that for mild cases, induction of measurable and significant antibody levels often takes several weeks (*9*). The antibody titers detected in both groups were initially lower and gradually increased to titers as high as 1:51,200 (**Figure 1C and D**). Although positive samples with lower antibody titers could constitute false positives, we believe that this again is a function of antibody kinetics, exemplified by the gradual increase in titers that can be clearly observed in the screening group (**Figure 1D**). Seropositive samples collected in the weeks ending February 23 and March 1 may potentially indicate that SARS-CoV-2 already had been introduced into the population of NYC earlier than initially detected. Overall, the titers in the sentinel group were significantly higher than in the screening group (**Figure 1F**), which is likely a function of disease severity in individuals in the sentinel group.

In order to determine which subgroup(s) were driving the rise in seroprevalence, we further separated the screening group into four subgroups: (i) “OB/GYN” visits and deliveries (n=1,366 samples); (ii) “Oncology” visits and treatment hospitalizations (n=1,319); (iii) “Surgery”, including various elective surgeries, transplant surgeries, pre-op medical assessments and related visits (n=544); and, (iv) “Cardiology”, including cardiology office visits and other regular office/treatment visits (n=183). This rise was mostly driven by “OB/GYN” visits and deliveries, which showed an early increase in seroprevalence in the week of March 29 (9.6%) followed by continued rise to 15.6% and 26.6% in the weeks ending April 12 and 19, respectively (**Figure 2A**). Seroprevalence in “Oncology” patients increased during the same time frame, but appears to plateau between 8-9% during these three weeks (**Figure 2B**). Similar seroprevalence trends were observed in the “Surgery” and “Cardiology” and other office visits subgroups, although the small number of specimens limited conclusions for these subgroups individually since confidence intervals were very wide (**Figure 2C and D**).

**Fig. 2.**
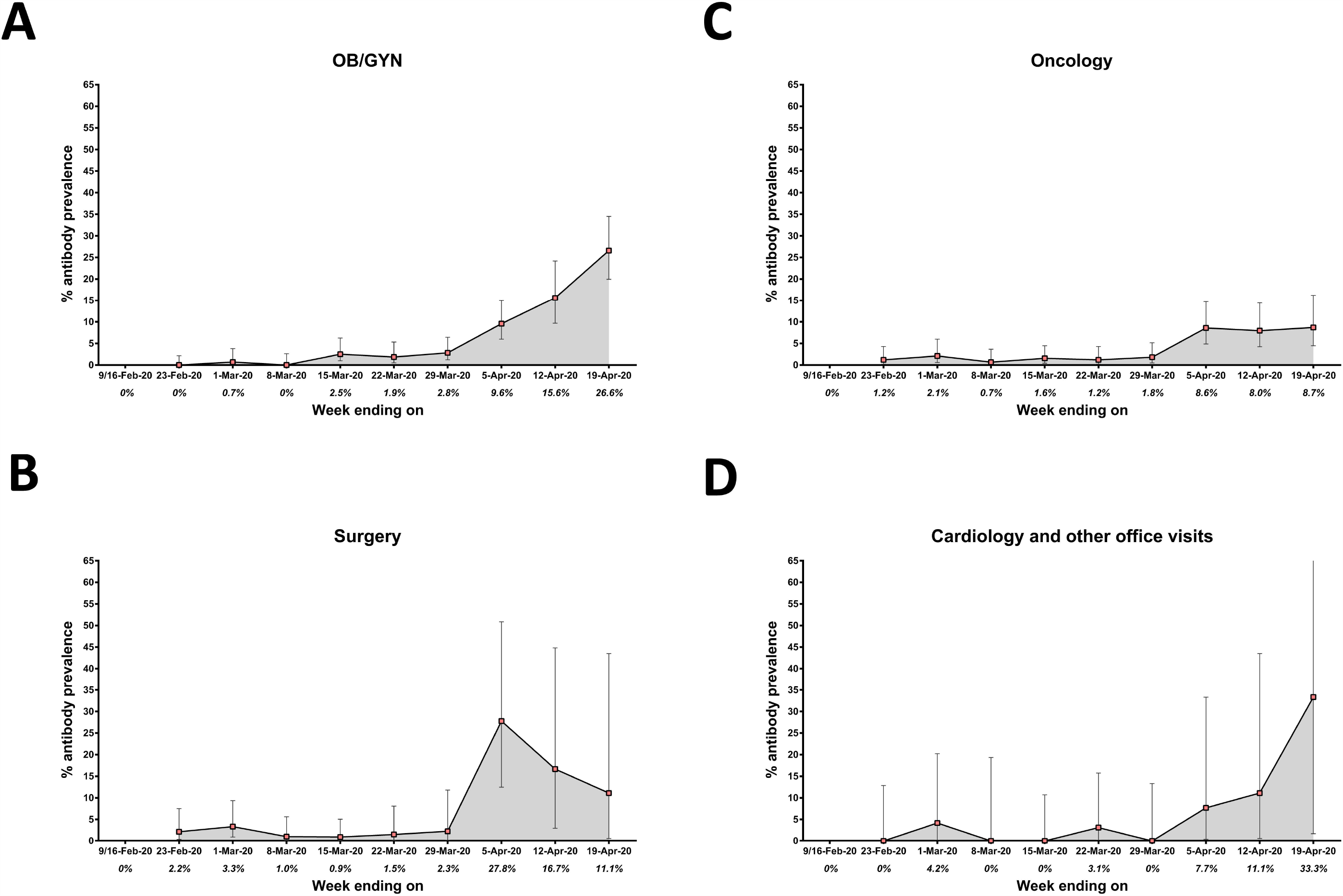
Seroprevalence in the different screening subclasses over time. Seroprevalence for OB/GYN **(A)**, surgery **(B)**, oncology **(C)** and cardiology and related office visits **(D)** groups in the weeks ending February 9 (first two weeks combined) to April 19, 2020.

Although our specimen sampling is biased and is not a true representation of the entire population, it nevertheless provides a window into the extent of seroprevalence in NYC. The sentinel samples are enriched in COVID-19 cases, since this group was designed to serve as a positive control. Due to the dramatic upsurge in SARS-CoV-2-related ED visits and admissions during the February-April 2020 time frame, as expected, a very high seroprevalence and a rapid increase in titers was detected in this group, consistent with its role as a positive control. In contrast, the screening group was enriched with individuals who would likely be cautious to avoid exposure to the virus, including pregnant women and patients with malignancies. The seroprevalence in the screening group consequently may be an underrepresentation of the seroprevalence in the general population. The 19.3% seroprevalence we found in this group for the week ending April 19 is consistent with a report from the New York–Presbyterian Allen Hospital and Columbia University Irving Medical Center that found 15.4% of pregnant women who delivered infants at their facilities between March 22 and April 4, 2020 were infected with SARS-CoV-2 and were mostly asymptomatic (*10*). This tracks well with seroprevalence in the screening group, which was between 10.1 and 19.3% in the weeks following April 5. A serosurvey conducted by the New York State Department of Health (NYSDOH) determined that between April 19 and 28, the seroprevalence for SARS-CoV-2 in the NYC metropolitan region was 22.7% (*11*), matching very well with the data for our screening samples from the week ending April 19. Of note, these numbers fall significantly below the threshold for community immunity, which has been estimated to require at least a seropositivity rate of 67% for SARS-CoV-2 (*4*). Based on the population of NYC (8.4 million), we estimate that by the week ending April 19, approximately 1,621,200 individuals had been infected with SARS-CoV-2. Taking into account the cumulative deaths in the city by April 19 (11,413), this suggests a preliminary IFR of 0.704%. This is in stark contrast to the IFR of the 2009 H1N1 pandemic which was estimated to be between 0.01% and 0.001% (*12*). We will continue to conduct this longitudinal serosurvey for at least one year, and expect that the seroprevalence for the screening group will slightly increase and then plateau for the time frame between the end of April and June, due to rapidly declining numbers of cases in NYC. If antibody titers remain stable, the seroprevalence would likely not change significantly unless new infections rise again or vaccines would become available.

## Data Availability

Data shown in the manuscript is available from the corresponding author on reasonable request.

## Acknowledgements

We thank Louella Rudon, Hanna Glazowski, and the team members of the Blood Bank and Rapid Response Laboratories of the Mount Sinai Health System for their assistance with acquisition of plasma samples. We are indebted to Dr. Carlos Cordon-Cardo for his continued support and encouragement. We would also like to thank the Technology Development Facility led by Dr. Robert Sebra for providing additional BSL2 space for sample collection. We are grateful for the continuous expert guidance provided by the ISMMS Program for the Protection of Human Subjects (PPHS).This work was partially supported by the NIAID Centers of Excellence for Influenza Research and Surveillance (CEIRS) contract HHSN272201400008C (FK, for reagent generation), Collaborative Influenza Vaccine Innovation Centers (CIVIC) contract 75N93019C00051 (FK, for reagent generation), and the generous support of the JPB foundation, the Open Philanthropy Project (#2020-215611) and other philanthropic donations.

## Supplementary Materials

**Suppl. Table 1:** Two-by-two contingency table to calculate sensitivity and specificity of SARS-CoV-2 antibody test.

|  |  | COVID19 |  |  |
| --- | --- | --- | --- | --- |
|  |  | positive | negative | total |
| Test | positive | 38 | 0 | 38 |
|  | negative | 2 | 74 | 76 |
|  | total | 40 | 74 | 114 |

|  | Value | 95% CI |
| --- | --- | --- |
| **Sensitivity** | 0.95 | 0.835 to 0.9911 |
| **Specificity** | 1 | 0.9507 to 1 |
| **Positive Predictive Value** | 1 | 0.9082 to 1 |
| **Negative Predictive Value** | 0.9737 | 0.909 to 0.9953 |

\*Wilson/Brown test to compute CIs

**Suppl. Table 2:**
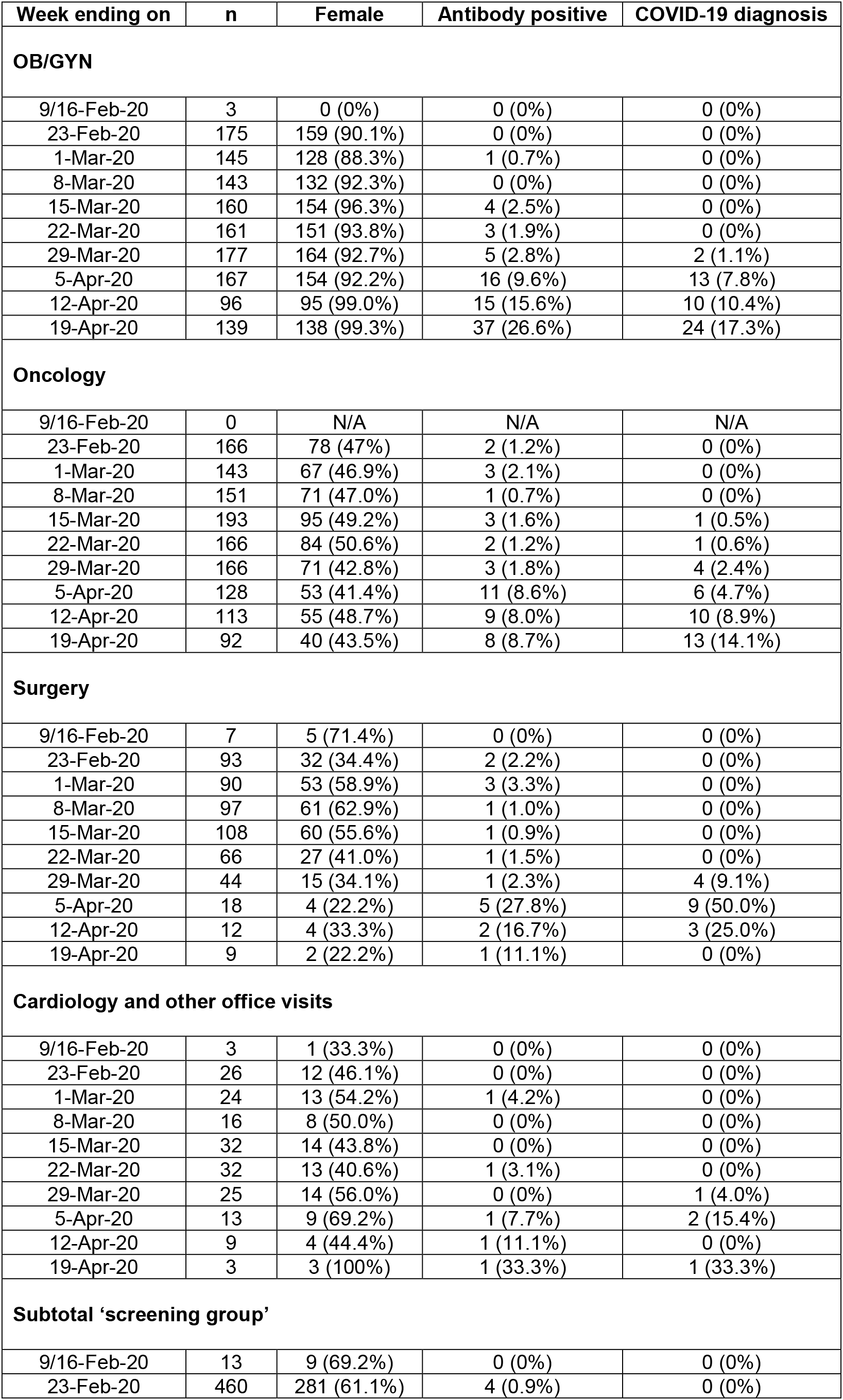
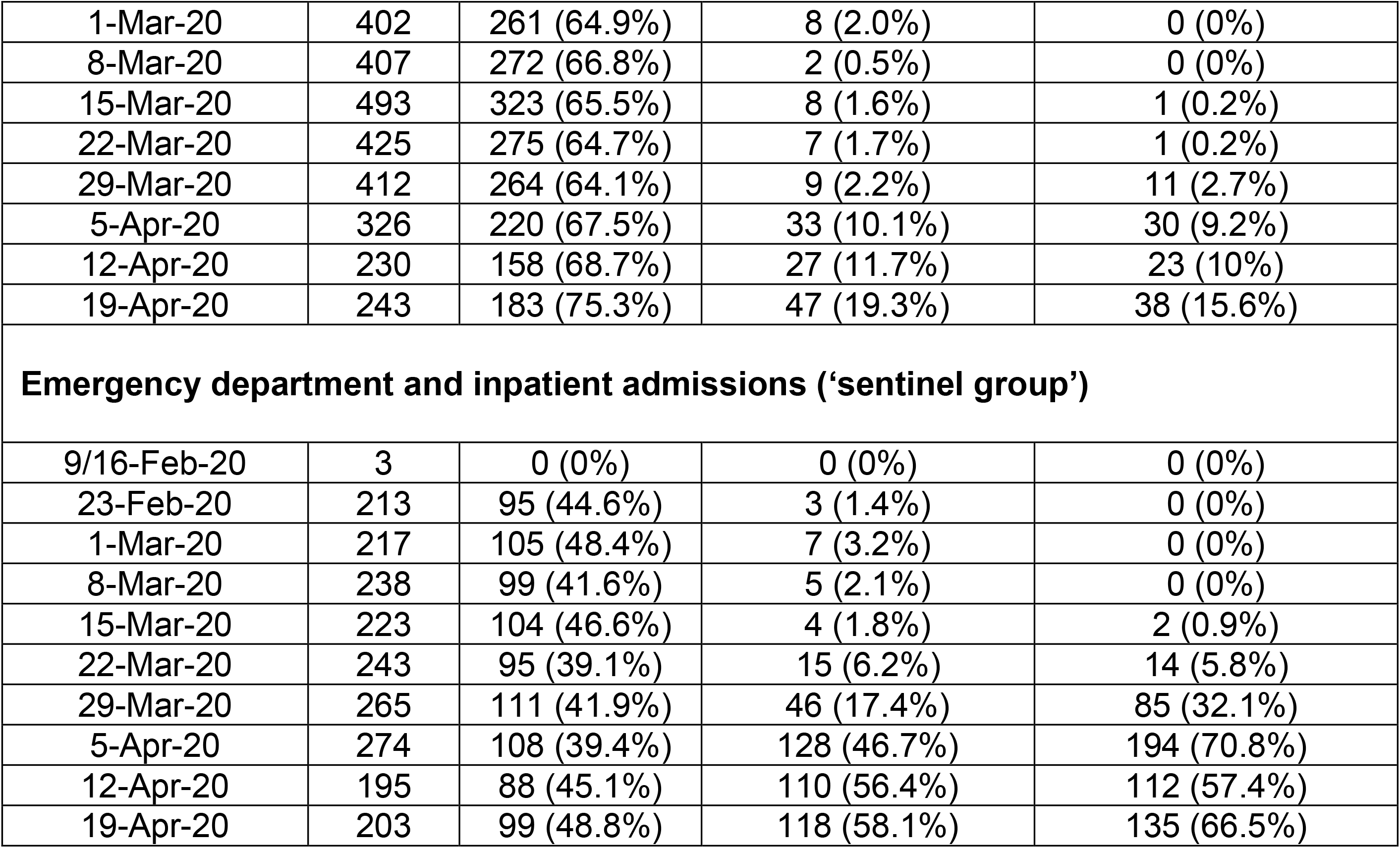
Detailed sample numbers and seroprevalence per week.

## Materials and Methods

### Study participants and human samples

Cross-sectional plasma samples that were stored for standard of care medical purposes by the MSHS Clinical Pathology laboratories were sampled beginning from the week ending February 9, 2020. About 230-460 plasma samples per week (starting from the week ending on February 23, 2020) were selected from the ‘screening group’ patient setting from patients that went for OB/GYN visits and deliveries, oncology visits and treatment hospitalizations, surgeries, transplant surgeries, pre-operative medical assessments and related visits as well as cardiology office visits and other regular office visits (see **Table S1** for detailed numbers and breakdown per cohort). About 200 plasma samples per week were selected from an inpatient cohort setting, consisting of plasma from patients that were admitted to the emergency department or to inpatient care (‘sentinel group’). Plasma samples were chosen in a blinded and unbiased manner. Longitudinal samples from the same patients were included in the analysis if the time points between sampling were at least seven days or more apart since this was seen as independent sampling of the population. For some individuals, a PCR test for viral RNA was performed to diagnose COVID-19 infection. The collection and testing of plasma was approved by the Mount Sinai Hospital Institutional Review Board, protocol HS# <u>20-03253</u>.

### Recombinant proteins

The recombinant RBD and spike protein of SARS-CoV-2 were generated and expressed as previously described (*7, 8*). In brief, the mammalian cell codon-optimized nucleotide sequences for RBD (amino acids 319-541) including a signal peptide and hexahistidine tag or the soluble version of the spike protein (amino acids 1-1,213) including a signal peptide, C-terminal thrombin cleavage site, T4 foldon trimerization domain and hexahistidine tag were cloned into the mammalian expression vector pCAGGS. The nucleotide sequence of the spike protein was additionally modified to remove the polybasic cleavage site and two stabilizing mutations were introduced. The expression plasmids are available at BEI Resources Repository (https://www.beiresources.org/).

Recombinant proteins were produced in Expi293F cells (Thermo Fisher) using the ExpiFectamine 293 Transfection Kit (Thermo Fisher) according to manufacturer’s instructions. Proteins were purified by gravity flow using Ni-NTA Agarose (Qiagen) and concentrated in Amicon centrifugal units (EMD Millipore). Purified proteins were analyzed by reducing sodium dodecyl sulfate– polyacrylamide gel electrophoresis (SDS-PAGE) and correct folding was confirmed by performing ELISAs with RBD-specific monoclonal antibody CR3022 (*13, 14*) or 2B3E5.

### Enzyme-linked immunosorbent assay (ELISA)

The serological assays were performed as previously described in detail following a two-step ELISA protocol (*7, 8*). In the first step, plasma samples were screened in a high-throughput assay using the recombinant RBD protein.

Ninety-six-well microtiter plates (Thermo Fisher) were coated with 50 µL recombinant RBD protein at a concentration of 2 µg/mL overnight at 4°C. The next day, the plates were washed three times with PBS (phosphate-buffered saline; Gibco) supplemented with 0.1% Tween-20 (T-PBS; Fisher Scientific) using an automatic plate washer (BioTek). The plates were blocked with 200 µL blocking solution consisting of PBS-T with 3% (w/v) milk powder (American Bio) and incubated for 1 h at room temperature. As a general safety precaution, plasma samples were heat inactivated for 1 h at 56°C. The blocking solution was thrown off the plates and 100 µL of plasma samples diluted 1:50 in PBS-T containing 1% (w/v) milk powder were added to respective wells of the microtiter plates. After 2 h the plates were washed three times with PBS-T and 50 µL anti-human IgG (Fab-specific) horseradish peroxidase antibody (HRP, produced in goat; Sigma, #A0293) diluted 1:3,000 in PBS-T containing 1% milk powder was added to all wells and incubated for 1 h at room temperature. The microtiter plates were washed three times with PBS-T and 100 µL SigmaFast *o*-phenylenediamine dihydrochloride (OPD; Sigma) was added to all wells. The reaction was stopped after 10 min with 50 µL per well 3M hydrochloric acid (Thermo Fisher) and the plates were read at a wavelength of 490 nm with a plate reader (BioTek). Plasma samples that exceeded an OD_490_ cutoff value of 0.15 were categorized as presumptive positives and were tested in a second step in confirmatory ELISAs using full-length, recombinant spike protein.

To perform the confirmatory ELISAs, the plates were coated and blocked as described above except full-length spike protein at a concentration of 2 µg/mL was added to the plates. After 1 h the blocking solution was removed, presumptive positive plasma samples serially diluted in 1% milk prepared in PBS-T were added and the plates incubated for 2 h at room temperature. The remainder of the assay was performed as described above. The data were analyzed in Microsoft Excel and GraphPad Prism 7. The cutoff value was set as an OD_490_ of 0.15 and true positive samples were defined as samples that exceeded an OD_490_ value of 0.15 at a 1:80 plasma dilution. The endpoint titer was calculated and defined as the last dilution before the signal dropped below an OD_490_ of 0.15. For samples that exceeded an OD_490_ of 0.15 at the last dilution (1:12,800 for samples of weeks ending on March 29 and April 5; 1:6,480 for samples of weeks April 12 and April 19), a four-parameter curve fit (variable slope) was applied and the endpoint titer determined by interpolation.

The sensitivity and specificity of the assay were determined using a panel of serum and/or plasma of 40 patients that had PCR-confirmed SARS-CoV-2 infection (true positives) and 74 negative control samples (56 samples that were taken before the pandemic and 18 samples without confirmed SARS-CoV-2 infection; true negatives). The PPV and NPV were determined taking into account the ratio of true positives and true negatives (seroprevalence of 35%) in the panel. Importantly, using the 100% specificity determined using the panel and assuming a low (e.g. 1%) true seroprevalence in the test group would not change the PPV.

### Statistical analysis

The 95% CI of the seroprevalence was calculated assuming binomial data based on methods by Wilson/Brown (*15*). Significant differences in endpoint titers between the sentinel and screening groups were identified by the Mann-Whitney U test. The 95% CI for assay sensitivity, specificity, positive predictive value and negative predictive value were determined using methods by Wilson/Brown.

